# Metabolomics differentiate cancer from non-cancer pleural effusions based on steroid lipids and acyl carnitines

**DOI:** 10.1101/2020.07.27.20162958

**Authors:** Katie Love, Ricardo MF da Costa, Manfred Beckmann, Anna Morley, Rahul Bhatnagar, Nick Maskell, Luis AJ Mur, Keir E Lewis

## Abstract

**Objective:** To identify and detect metabolomic markers to differentiate cancer related from other pleural effusions.

**Material and methods:** Un-targeted flow infusion electrospray mass spectrometry was used on a cohort of 100 samples from benign and malignant pleural effusions (12 primary lung cancers, 14 mesotheliomas, 24 other cancers, 25 congestive cardiac failure, 22 parapneumonic effusion, 3 empyema). Standard metabolomic statistical models for analysis were performed.

**Results:** Five novel markers were identified using univariate and multivariate receiving operator curve analyses using five discriminant features yielding a diagnostic accuracy of above 0.91 (95% CI: 0.839-0.976) with a sensitivity of 0.97 (95% CI 0.952-0.979) and specificity 0.87 (95% CI 0.852-0.894).

**Conclusion:** Individual biomarkers with the highest accuracy (>0.84) were all raised in cancer and are involved in metabolism of sterol-lipids and acyl carnitines linked to β-oxidation. Our markers performed better than those previously published. Further, these published markers failed to achieve their stated accuracies in our data.

## Introduction

Accumulation of pleural fluid is a common pathology in over 50 recognised aetiologies ranging from treatable infections, to malignant mesothelioma [1]. For more than 40 years ‘Light’s criteria’ has been the gold standard for the diagnosis of pleural effusions, able to differentiate effusions with a diagnostic accuracy of 93-96% [2]. However, they are known to mis-classify some transudates as exudates e.g. especially in patients using diuretics [3], which may have significant implications for clinical management. Further, cytological confirmatory tests for malignancy through pleural aspiration account for an accuracy of only 60 %, with a sensitivity of 40-87 % [4]. Better diagnostics can be developed with new technologies, such as metabolomics, and have yielded accurate, reliable, and sensitive tests for lung cancer using other non-invasive samples such as saliva and urine [5]–[7].

The metabolome describes the large numbers of metabolites within a given sample. Untargeted metabolomic profiling, aims to comprehensively detect, quantify, and identify metabolites in a given sample. Flow Infusion Electrospray-Mass Spectrometry (FIE-MS) is one technique that allows high-throughput and highly sensitive detection of metabolites. Metabolomics increases the fundamental understanding of biochemical pathways in normal and abnormal pathologies.

Although metabolomics could be of major benefit to clinical medicine [8], it has yet to deliver on this potential. Despite having a comprehensive list of processing and reporting standards for metabolomic studies, many published papers fail to adhere to them – resulting in a lack of reproducible findings and a scarcity of translating metabolomics into clinical practice [9].

Herein we use FIE-MS to generate metabolomic data from cancerous and non-cancerous pleural effusions and compare its accuracy with standard approaches including Lights criteria and other published pleural fluid biomarkers. We also aim to use our data set as a rudimentary external validation cohort for previously published biomarker studies.

## Methods

In this pilot, proof of concept study, 100 pleural effusion samples were selected from a UK, cohort-based repository. Samples were obtained from patients attending a tertiary clinical centre between 2015-2018. A cancer diagnosis was confirmed cytologically or histologically and non-cancer diagnosis confirmed either cytologically or histologically from the pleural space, in line with usual clinical standards.

Samples were chosen to consist of an equal number of 50 cancers (12 primary (LC), 14 mesothelioma (MPM) 24 other secondary cancers), and 50 samples from non-cancer (25 congestive cardiac failure (CCF), 22 Complicated Parapneumonic Effusion (CPPE) and 3 with empyema).

18 patients were diagnosed on their first pleural tap (x4 LC, 1 mesothelioma, 13 other cancer), with 32 samples obtained from further tests (CT-guided biopsy, video assisted thoracoscopy, repeat pleural taps, surgery). Patient metadata are summarised in Supplementary Table 1.

### Processing of pleural effusions

All samples were processed for metabolomics in a blinded manner. A two-step extraction was used for metabolomic analysis. Aliquots were thawed on ice then vortexed, followed by centrifugation (1800 ×*g* for 10 min). 100 μL of supernatant was added to 50mg of acetone-washed glass beads (≤106μm), with 1mL of 4:1 (v/v) mix of MeOH:chloroform (HPLC grade). Samples were vortexed, shaken (60 min at 4°C), left to precipitate (−80°C for 20 min), then centrifuged (1800 ×*g* for 10 min). 100 μL of the upper layer was re-extracted following the same procedure.

### Flow infusion electrospray mass spectrometry (FIE-MS)

60 μL of the supernatant was transferred to a glass vial with insert. Sample injection order was randomised to reduce batch effects and run on an Exactive™ Orbitrap Mass Spectrometer (Thermo Scientific). Data were acquired in alternating positive and negative ionisation modes over 4 scan ranges (15–110, 100–220, 210–510, and 500–1200 *m/z*) 5 min acquisition time. Following the infusion of each sample, mass spectra were derived from the mean of 20 scans about the apex of the peak signal, Data were normalised against the total ion count and the intensity value for each individual *m/z* (ranging from 55.93187 to 1178.146 Da) and expressed as a percentage of the total ion count. Data are given in Supplementary Table 2.

### Data and statistical analysis

Disease classifications were known to the data analyst, and missing values were assigned as ‘NA’. Data were Bonferroni corrected for multiple comparisons. Principal Component Analyses (PCA), Partial Least Square Discriminant Analysis (PLS-DA), ANOVA, Receiver Operating characteristic (ROC), and Area Under Curve (AUC) analyses were applied using the MetaboAnalyst 4.0 platform [10]. Our ROC curves were generated by Monte-Carlo cross validation using subsampling (2/3 to evaluate feature importance, and 1/3 to validate the models). ROC-AUC analyses were conducted on metabolites with a Variable Importance in Projection (VIP) score of >2. The 5 metabolites were chosen based on VIP scores (>2) and statistical significance (*P*<0.01) (corrected for false discovery rates). Metabolites were identified based on PubChem [https://pubchem.ncbi.nlm.nih.gov/], HMDB [http://www.hmdb.ca/], KEGG [https://www.genome.jp/kegg/compound/] and CheBI databases [https://www.ebi.ac.uk/chebi/] which were interrogated using MZedDB [http://maltese.dbs.aber.ac.uk:8888/hrmet/index.html] and DimeDB [https://dimedb.ibers.aber.ac.uk/] interfaces.

A literature search was conducted via PubMed, into pleural effusion biomarkers using a variety of analytical platforms. 7 studies which reported sufficient details to identify metabolites were included in our analyses [11]–[17].

## Results

PCA show that patients clustered between the two broad primary pathologies of cancer and non-cancer (Fig. 1A). The major sources of variation were investigated, and 5 metabolites increased in cancer with AUC accuracies of 0.914 (95% CI: 0.839-0.976) with a sensitivity of 0.97 (95% CI 0.952-0.979) and specificity 0.87 (95% CI 0.852-0.894). (Fig. 1B). These metabolites were lipid steroids; hydroxyprogesterone (AUC 0.85, 95% CI: 0.767-0,913), stigmastentriol (AUC 0.838, 95% CI: 0.773-0.901), hydroxycholesterol (AUC 0.84, 95% CI: 0.773-0.897) and acylcarnitines; hexadec-2-enoyl carnitine (AUC 0.83, 95% CI: 0.739-0.898), hydroxyhexadecanoylcarnitine (AUC 0.81, 95% CI: 0.726-0.873).

**Figure 1.**
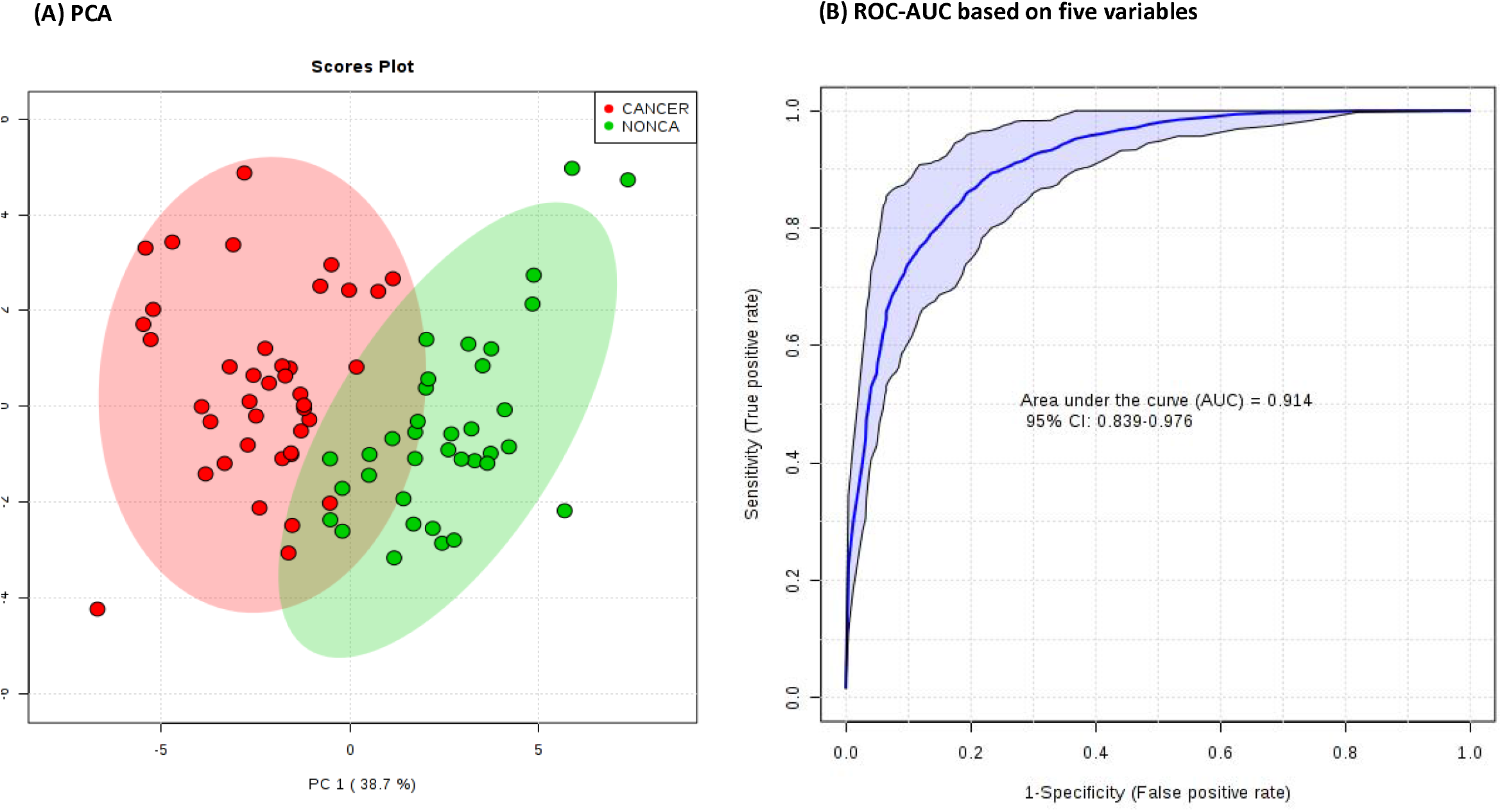
Metabolomics can distinguish between malignant and non-malignant causes of pleural effusions. **(A)** Principal Component Analysis (PCA) the larger circles in the PCA represent 95% CI for each sample class) and **(B)** multivariate ROC curve comparison evaluated the performance of CANCER and non-cancer (NON) samples

Due to limitations in using multivariate models with relatively small sample sizes, we employed another multivariate classifier approach, Random Forest (RF). This confusion matrix showed a class error for the cancer group of only 0.12 and for non-cancer of only 0.03. The out-of-bag, error was used as a test sample to obtain an unbiased estimate of the classification error, which was only 0.066.

Our data was interrogated for previously published pleural fluid biomarkers. These were identified on accurate mass (5 ppm) and predicted positive and negative ionisation patterns. Where metabolites could also be found in our data, pairwise comparisons were conducted, and AUC values were generated (Supplementary Figure 1). Univariate or multivariate assessments of these previously published biomarkers did not perform as well as the five that we targeted in our data.

## Discussion

This study demonstrates that metabolomic approaches can be applied to stored pleural fluid to differentiate malignancies from non-cancer aetiologies, with a high sensitivity and specificity of 97% and 87% respectively. No other study has reported untargeted FIE-MS to pleural fluid. Due to the exploratory nature of this pilot study, no external validation set was available. However, our data was used as a validation set for previously published pleural fluid biomarkers.

The metabolites we discovered included hydroxyprogesterone, stigmastentriol, and hydroxycholesterol. These indicate an upregulation of lipid-related metabolic pathways in pleural effusions caused by cancer. This was further indicated by the acyl carnitines which is the form that long-chain fatty acids take as they cross the inner mitochondrial membrane for subsequent β-oxidation [18]. Increased levels of these metabolites are consistent with the increased fatty acid synthase expression which occurs during carcinogenesis [19]. Further, our results compliment the findings of Ziananro et al., who linked the differentiation of cancer from non-cancer pleural effusions using proton magnetic resonance (^1^H-NMR) techniques, to increased lipid metabolism, along with increased expression of branched amino acids and lactate metabolism [17]. Che et al., also applied metabolomics to pleural fluid using metabolomics to differentiate the underlying causes of pleural fluid accumulation. Their study focused on tuberculous versus cancer effusions, achieving a sensitivity of 93% and specificity of 86% [16]. Both our studies indicate that metabolomics offers a potential real-world benefit in the diagnosis and differentiation of pleural effusion cause.

Other genetic and protein biomarkers have been reported to differentiate the cause of pleural effusions but very few are accurate enough to be accepted into clinical practice yet. Metabolomics does offer potential to identify a suite of new potential biomarkers. Our biomarkers appear to be more accurate than previously reported biomarkers for differentiating between Cancer and Non-cancer causes (Table 1). However, this observation highlights the requirement for a larger, multisite, blinded study to validate all the pleural biomarkers that have been defined to date.

**Table 1:** Assessment of putative PE biomarkers from the literature

| Name | AUC | T-test (*P*) | Log<sub>2</sub> FC | Reference |
| --- | --- | --- | --- | --- |
| R-2-Hydroxyhexadecanoic acid | 0.7313 | 7.52E-10 | 0.77869 | Ho et al., (2016) |
| Choline | 0.7162 | 4.17E-08 | 0.73284 | Wang et al., (2017) |
| Oleic acid | 0.679 | 2.16E-06 | 0.75869 | Lam & Law (2014) |
| Linoleic acid | 0.6742 | 6.38E-06 | 0.79921 | Lam & Law (2014) |
| Ceramide (d18:1/16:0) | 0.6718 | 0.002699 | 0.44135 | Lam & Law (2014) |
| Docosapentaenoic acid | 0.6714 | 6.29E-04 | 0.59109 | Ho et al., (2016) |
| Eicosatrienoic acid | 0.6635 | 0.00262 | 0.49347 | Ho et al., (2016) |
| Ornithine | 0.6623 | 2.24E-04 | -0.56498 | Wang et al., (2017) |
| Linolenic acid | 0.6546 | 4.54E-05 | 0.52851 | Lam & Law (2014) |
| Tetradecadienoic acid | 0.6431 | 0.001177 | 0.46765 | Ho et al., (2016) |
| LysoPEtn(P16:0) | 0.6376 | 0.001096 | 0.78928 | Ho et al., (2016) |
| Eicosapentaenoic acid | 0.6253 | 0.026597 | 0.3196 | Ho et al., (2016) |
| Adrenic acid | 0.6151 | 0.11403 | 0.32006 | Ho et al., (2016) |
| Tyrosine | 0.596 | 0.013926 | -0.38366 | Wang et al., (2017) |
| SM(d42:2) | 0.59435 | 0.011886 | -0.50853 | Ho et al., (2016) |
| Creatinine | 0.5898 | 0.071719 | 0.58439 | Wang et al., (2017) |

In conclusion, we have further demonstrated the promise of untargeted metabolomic based discovery of biomarkers, for differentiating cancer vs non-cancer pleural effusions. We report the importance of lipid biomarkers and fatty acid synthase expression. The increased lipid turnover is biologically plausible and reported in cancer metabolism. However, future validation on larger and different clinical sets is required.

## Data Availability

data are uploaded as metadata and are freely available

## Abbreviations

CCF: Congestive Cardiac Failure
COPD: Chronic Obstructive Pulmonary Disease
CPPE: Complicated Parapneumonic Effusion
FIE-MS: Flow Infusion Electrospray ion Mass Spectrometry
IPF: Idiopathic Pulmonary Fibrosis
PCA: Principal Component Analyses
ROC: Receiver Operating Characteristic
AUC: Area under the Curve
VIP: Variable Importance in Projection
RF: Random Forest

## Acknowledgments

We appreciate the technical support provided by Helen Phillips with mass spectrometry (Aberystwyth University, UK).

## Financial support

RdC was funded by a Tenovus iGrant TIG2016-26. KL is supported by a Knowledge Economy Skills Scholarship (KESS 2) and further supported by European Social Funds (ESF).

## Notes

Supported by grants from Knowledge Economy Skills Scholarships (KESS) and TENOVUS Cancer Charity UK. Funders had no involvement in the study design, collection, analysis or interpretation of data. Nor in the writing of the report, or in the decision to submit the article for publication.

### Competing Interest Statement

The authors have declared no competing interest.

### Clinical Trial

this was a prospective study based on biobanked samples.

### Author Declarations

National Health Service (UK); Health Research Authority https://www.hra.nhs.uk/planning-and-improving-research/application-summaries/research-summaries/novel-technologies-for-diagnosing-and-monitoring-pulmonary-diseases/

